# Mental Health of HBCU College Students during the COVID-19 Pandemic

**DOI:** 10.1101/2021.07.22.21260878

**Authors:** Sharron Xuanren Wang, Jarid Goodman

## Abstract

**Objective:** This study investigated rates and predictors of mental health issues (e.g., depression and anxiety) in a sample of college students currently attending a historically Black college/university (HBCU) during the COVID-19 pandemic.

**Participants/Methods:** 98 undergraduate students (81 female and 17 male) completed an online survey containing questions about demographics, socioeconomic status, academic characteristics, and pandemic-related concerns. The survey also included PHQ-9 and GAD-7 questionnaires to evaluate depression and anxiety, respectively.

**Results:** 49% of students met the clinical cutoff for depression, 39% for anxiety, and 52% for depression and/or anxiety. Significant predictors of meeting the cutoffs included parental job loss/hour reduction, being a senior, and feeling that the pandemic negatively impacted daily life, among other factors. Demographic variables (age, gender, etc.) had no effect.

**Conclusion:** HBCU students show high rates of depression and anxiety during the COVID-19 pandemic, which may be predicted based on the student’s academic, socioeconomic, and pandemic-related concerns.

## INTRODUCTION AND BACKGROUND

The coronavirus disease-2019 (COVID-19) pandemic has introduced a wide range of economic and health-related challenges. Accumulating evidence suggests that financial and economic concerns, in addition to negative health-related events such as COVID-19 infection or the loss of family members to the disease, may have caused a surge in mental health issues around the world (Marroquín, Vine, & Morgan, 2020; Rajkumar, 2020; White & Van Der Boor, 2020; Xiong et al., 2020). In the United States, groups facing heightened mental health issues during the pandemic include college students (Huckins, et al., 2020; Lei, Huang, Zhang, Yang, Yang, & Xu, 2020) and racial and ethnic minorities (Czeisler et al., 2020). The present study was designed to gain a deeper understanding of the factors potentially influencing mental health among college students attending a historically Black public university (HBCU) in the Northeastern U.S. during the COVID-19 pandemic.

College students in general face high rates of mental health problems, relative to the general population, as a result of concerns about academic performance, finances, being away from home and family, new social relationships, and plans after college, among other concerns (Aselton, 2012; Beiter et al., 2015). Evidence has indicated a steady rise in mental health concerns over the past 20 years for college students, such as increasing rates of anxiety and depression (American College Health Association, 2018). The COVID-19 pandemic has introduced new challenges with the potential to further impact the mental health of college students, such as campus closures, remote learning, isolation from friends, parental job loss, concerns about financing their education, and uncertainty about finding a job after graduation in a labor market ravaged by the pandemic. In addition, students might also harbor concerns about their health and the health of immediate family members, especially family members of advanced age or those with pre-existing health conditions that increase the risk of negative health outcomes following COVID-19 infection. Indeed, a recent national survey of college students reported increased rates of anxiety and depression during the COVID-19 pandemic (e.g., Healthy Minds Network and American College Health Association, 2020).

In addition to young adults and college students, there is accumulating evidence that the COVID-19 pandemic has had a disproportionately negative impact on racial and ethnic minorities in the Unites States (Goodman et al., 2020; Kujawa, Green, Compas, Dickey, & Pegg, 2020; Oppel Jr, Gebeloff, Lai, Wright, & Smith, 2020; Sáenz & Sparks, 2020). Black and Hispanic Americans have faced much higher rates of COVID-19 infection and death, compared to non-Hispanic Whites (Oppel Jr, Gebeloff, Lai, Wright, & Smith, 2020). In addition, unemployment rates have been higher and job recovery has proven relatively slower for African Americans and Hispanic Americans during the COVID-19 pandemic, compared to non-Hispanic Whites (Sáenz & Sparks, 2020). While some research has also shown increased rates of mental health problems for racial and ethnic minorities during the pandemic (Czeisler et al., 2020), no published work to our knowledge has examined what factors potentially modulate the negative mental health impact of the pandemic among college students of predominantly minority racial/ethnic backgrounds.

The present study employed an online survey that was distributed among undergraduate college students currently attending an HBCU in the Northeastern U.S. during the COVID-19 pandemic. The survey included brief clinical assessments for depression and anxiety, while also containing questions about demographics and other factors that potentially mediate mental health outcomes among college students during the pandemic, such as academic concerns and parental unemployment. The present study represents an important preliminary step toward understanding the factors affecting the mental health impacts of the COVID-19 pandemic for HBCU students.

## DATA AND METHODS

A survey was created using SurveyMonkey. The survey contained questions pertaining to demographic information, socioeconomic status, education, and experiences and concerns related to the COVID-19 pandemic. In addition, the survey used the Generalized Anxiety Disorder questionnaire – 7 item (GAD-7) and the Patient Health Questionnaire – 9 item (PHQ-9) to evaluate anxiety and depression, respectively. A link to the survey was distributed via email to undergraduate students at a public HBCU in a Northeastern state on May 17^th^ 2020. The university closed the campus in mid-March due to pandemic-related concerns, and all courses transitioned to online/remote instruction for the remainder of the Spring 2020 semester. All procedures were approved by the Institutional Review Board at Delaware State University. Upon completion of the survey, each respondent received $15 compensation. The survey closed on May 22^nd^. In total, 123 students completed the survey; however, 25 cases did not complete the mental health questionnaires and were therefore dropped from the study using list-wise deletion. The remaining respondents (N = 98) were included in the data analyses.

### Outcome Variables

The GAD-7 and PHQ-9 were used as mental health screens for anxiety and depression, respectively. Both scales demonstrate high reliability and validity among the general population of the United States, including for U.S. college students (Löwe et al., 2008; Kroenke, Spitzer, & Williams, 2001). Each questionnaire contains a list of symptoms, and the participant is instructed to rate how often they experienced each symptom within the past two weeks (0 = Not at all, 1 = Several days, 2 = More than half the days, 3 = Nearly every day). In the clinical setting, a score of 10 or greater on either questionnaire represents a cutoff point, suggesting the respondent should be further evaluated for anxiety or depressive disorders, respectively. This cutoff point is based on prior literature indicating a PHQ-9 score ≥ 10 shows 88% sensitivity and 88% specificity for major depression (Kroenke, Spitzer, & Williams, 2001). Likewise, a GAD-7 score ≥ 10 shows 89% sensitivity and 82% specificity for generalized anxiety disorder (Spitzer, Kroenke, Williams, & Löwe, 2006). In the present study, we employed the two questionnaires to create three binary outcome variables to measure whether each respondent met the GAD-7 cutoff (yes = 1; no=0), the PHQ-9 cutoff (yes = 1; no=0), and whether they met the GAD-7 and/or the PHQ-9 cutoff (yes = 1; no=0).

### Independent Variables

Independent variables for the present study included potential risk factors for mental health issues among college students during the COVID-19 pandemic. We used variables that past research has suggested may impact mental health (i.e., demographic, socioeconomic, health, and academic) and included additional variables potentially relevant during the pandemic. Specifically, demographic characteristics included age as a continuous variable, sex (female = 1, male =0), race and ethnicity (i.e., Hispanic black, non-Hispanic black, and others), and place of birth (i.e., native born and foreign born). Socioeconomic and health characteristics included father with Bachelor’s degree or higher (yes =1, no = 0), mother with Bachelor’s degree or higher (yes = 1, no=0), whether student was eligible for a Pell grant (yes = 1, no =0), whether at least one parent’s work hours were reduced at any point during the pandemic (yes = 1, no=0), whether at least one parent lost their job at any point during the pandemic (yes = 1, no=0), whether the student was currently working (yes = 1, no = 0), number of people living in the household as a continuous variable, and past diagnosis of a mental health problem (yes = 1, no = 0). Academic characteristics included college year (i.e., freshman, sophomore, junior, and senior or higher), major (i.e., STEM, health science, and others), and whether cumulative college GPA was 3.5 or higher (yes = 1, no = 0). Pandemic-related questions were all binary variables (yes = 1, no = 0) and included the following: whether the pandemic made them anxious about running out of food, water, and/or other essential items; whether the pandemic made them anxious about coursework; whether the pandemic made them forgetful; whether the pandemic negatively impacted their mental health; whether the pandemic negatively impacted their daily life; and whether the pandemic negatively impacted their academic performance (e.g., in terms of grades, completing assignments on time, etc.).

## Methods

Three logistic regression models were performed to identify risk factors for mental health issues among HBCU college students. The dependent variables were three binary variables, including meeting the cutoff for anxiety, meeting the cutoff for depression, and meeting the cutoff for depression and/or anxiety (for all variables: yes = 1, no=0, for all variables). The independent variables were potential risk factors, including demographic, socioeconomic/mental health, and academic characteristics, as well as pandemic-related information. All analyses were performed using STATA. The variance inflation factors (VIF) were calculated to diagnose multicollinearity.

## RESULTS

Table 1 displays descriptive statistics for the sample. The mean age of our sample was 20.1 (SD=1.53). There were about 83% students who identified as female and 17% identified as male. The vast majority of students identified as Black/African-American. Based on their ethnic identification, we further categorized Black students into Hispanic Black and non-Hispanic Black. About 8% of students were categorized as Hispanic Black, 83% as non-Hispanic Black, and 8% as Other. The category of Other included White, Asian, Native Americans, Hawaiian, and those who self-identified their race as “other”. Ninety-two percent of the sample were native-born and 8% were foreign-born. The demographic statistics of the sample is consistent with the university’s demographic profile. About 23% of the students reported their father had at least a Bachelor’s degree and about 52% reported their mother had at least a Bachelor’s degree. Approximately half (54%) indicated they were eligible for a federal Pell grant. Regarding parents’ employment during the pandemic, 46% of the students reported that at least one of their parents experienced a cut in hours at some point during the pandemic, and 24% of students reported that at least one of their parents lost their job at some point during the pandemic. In addition, 31% of the students were currently working either full-time or part-time. The mean number of people residing in the student’s household was 4.4 (SD=1.72). Notably, 11% of students reported having been previously diagnosed with a mental health disorder.

**Table 1.**
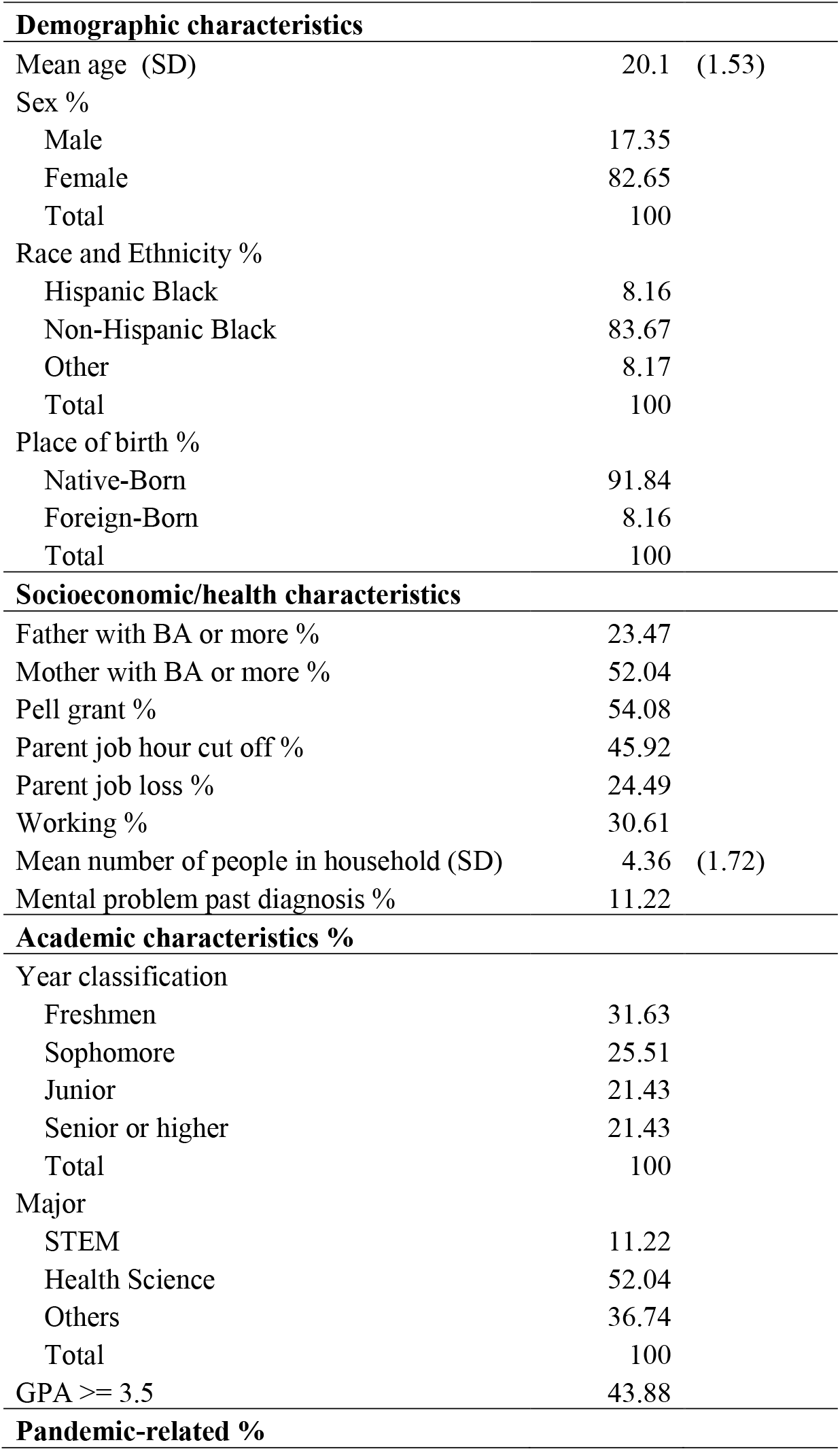
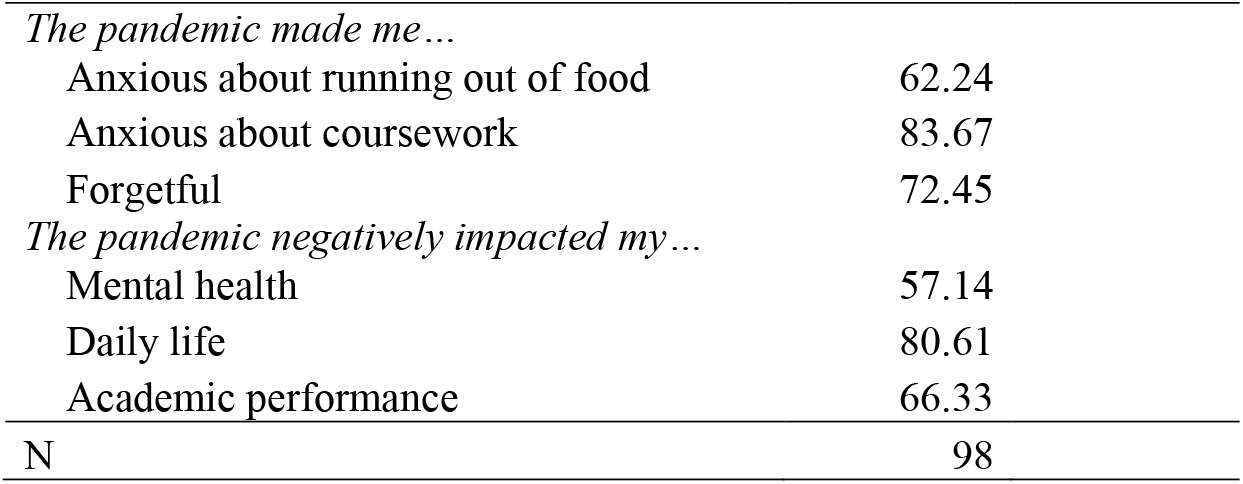
Descriptive Statistics

| **Demographic characteristics** |  |  |
| --- | --- | --- |
| Mean age (SD) | 20.1 | (1.53) |
| Sex % |  |  |
| Male | 17.35 |  |
| Female | 82.65 |  |
| Total | 100 |  |
| Race and Ethnicity % |  |  |
| Hispanic Black | 8.16 |  |
| Non-Hispanic Black | 83.67 |  |
| Other | 8.17 |  |
| Total | 100 |  |
| Place of birth % |  |  |
| Native-Born | 91.84 |  |
| Foreign-Born | 8.16 |  |
| Total | 100 |  |
| **Socioeconomic/health characteristics** |  |  |
| Father with BA or more % | 23.47 |  |
| Mother with BA or more % | 52.04 |  |
| Pell grant % | 54.08 |  |
| Parent job hour cut off % | 45.92 |  |
| Parent job loss % | 24.49 |  |
| Working % | 30.61 |  |
| Mean number of people in household (SD) | 4.36 | (1.72) |
| Mental problem past diagnosis % | 11.22 |  |
| **Academic characteristics %** |  |  |
| Year classification |  |  |
| Freshmen | 31.63 |  |
| Sophomore | 25.51 |  |
| Junior | 21.43 |  |
| Senior or higher | 21.43 |  |
| Total | 100 |  |
| Major |  |  |
| STEM | 11.22 |  |
| Health Science | 52.04 |  |
| Others | 36.74 |  |
| Total | 100 |  |
| GPA >= 3.5 | 43.88 |  |
| **Pandemic-related %** |  |  |

|  |  |
| --- | --- |
| *The pandemic made me...* |  |
| Anxious about running out of food | 62.24 |
| Anxious about coursework | 83.67 |
| Forgetful | 72.45 |
| *The pandemic negatively impacted my...* |  |
| Mental health | 57.14 |
| Daily life | 80.61 |
| Academic performance | 66.33 |
| N | 98 |

Regarding academic characteristics, 21% of students identified as a senior or higher. Moreover, 11% of students were majoring in a STEM field, and 52% were majoring in a health sciences field. Forty-four percent of students reported a cumulative GPA of 3.5 or higher. In response to the pandemic-related variables, 62% of students reported that the pandemic made them anxious about running out of food, water, and/or other essential items; 84% reported the pandemic made them more anxious about coursework; 72% reported the pandemic made them more forgetful; 57% reported the pandemic negatively affected their mental health; 81% reported the pandemic negatively affected their daily life; and 66% reported that the pandemic negatively affected their academic performance.

Table 2 summarizes mental health outcomes in our sample. The mean PHQ-9 score for the sample was 11.03 (SD = 7.16), and the mean GAD-7 score was 8.41 (SD=6.6). Notably, 49% of our sample met the clinical cutoff for depression (PHQ-9 ≥ 10), and 39% met the clinical cutoff for anxiety (GAD-7 ≥ 10). Moreover, about half (52%) met the cutoff for depression and/or anxiety. These findings indicate remarkably high rates of mental health issues in our sample, and that many students may meet full criteria for diagnosis of an anxiety disorder or depressive disorder upon further evaluation.

**Table 2.**
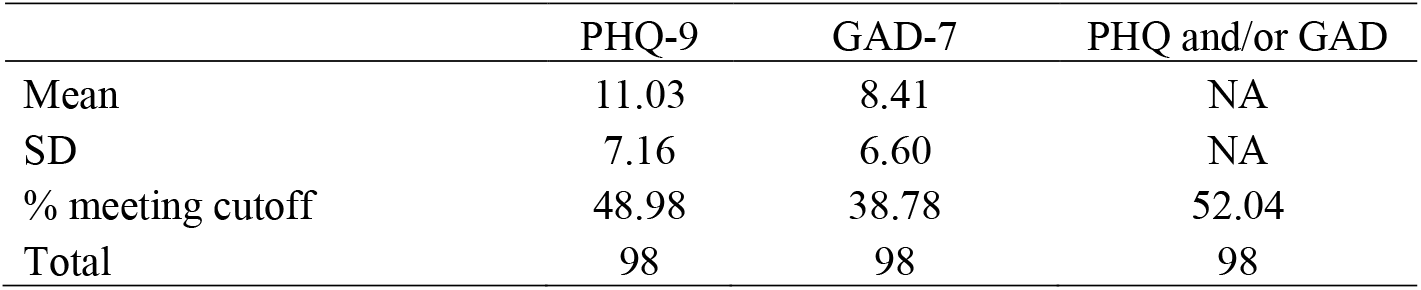
Mental Health Outcomes

Table 3 displays the results of three logistic regression models that test potential predictors of mental health outcomes among HBCU students. Odds ratios, standard errors, and p values are presented. None of the demographic characteristics that were tested showed a significant effect, indicating that demographic factors (e.g., race, gender, etc.) did not have a significant impact on depression and/or anxiety. In contrast, some predictors in the socioeconomic/mental health category demonstrated significant effects. Having at least one parent with reduced work hours during the pandemic served as a significant predictor of depression, anxiety, and depression and/or anxiety. For example, the odds ratio for parental cut in work hours in the PHQ-9 model was 8.16 (p=0.01), indicating that the odds of meeting the PHQ-9 cutoff for students whose parent(s) experienced a cut in work hours was 8.16 times larger than the odds of meeting the PHQ-9 cutoff for the students whose parent(s) did not experience a cut in hours, holding all other variables constant. The odds ratio for parental job loss was also significant in the GAD-7 model (OR=7.31, p= 0.052), indicating that the odds of meeting the GAD-7 cutoff was predicted to be 7.31 times larger for students whose parent(s) experienced job loss during the pandemic than those whose parents did not experience job loss, holding other variables constant. The number of people living in the student’s household also has a significant effect on the odds of meeting the GAD-7 cutoff (OR=1.57, p =0.057) as well as the odds of meeting either the PHQ-9 and/or GAD-7 cutoff (OR=1.55, p = 0.051). For example, if the number of people living in the household increases by 1, the odds of meeting the GAD-7 cutoff are predicted to increase by 57%, and the odds of meeting either the GAD-7 and/or the PHQ-9 cutoff are predicted to increase by 55%. Lastly, having received a past diagnosis of a mental health disorder significantly predicted whether a student met the GAD-7 cutoff (OR=7.94, p=0.082), as well as the PHQ-9 and/or GAD-7 cutoff(s) (OR = 15.91, p = 0.026).

**Table 3.**
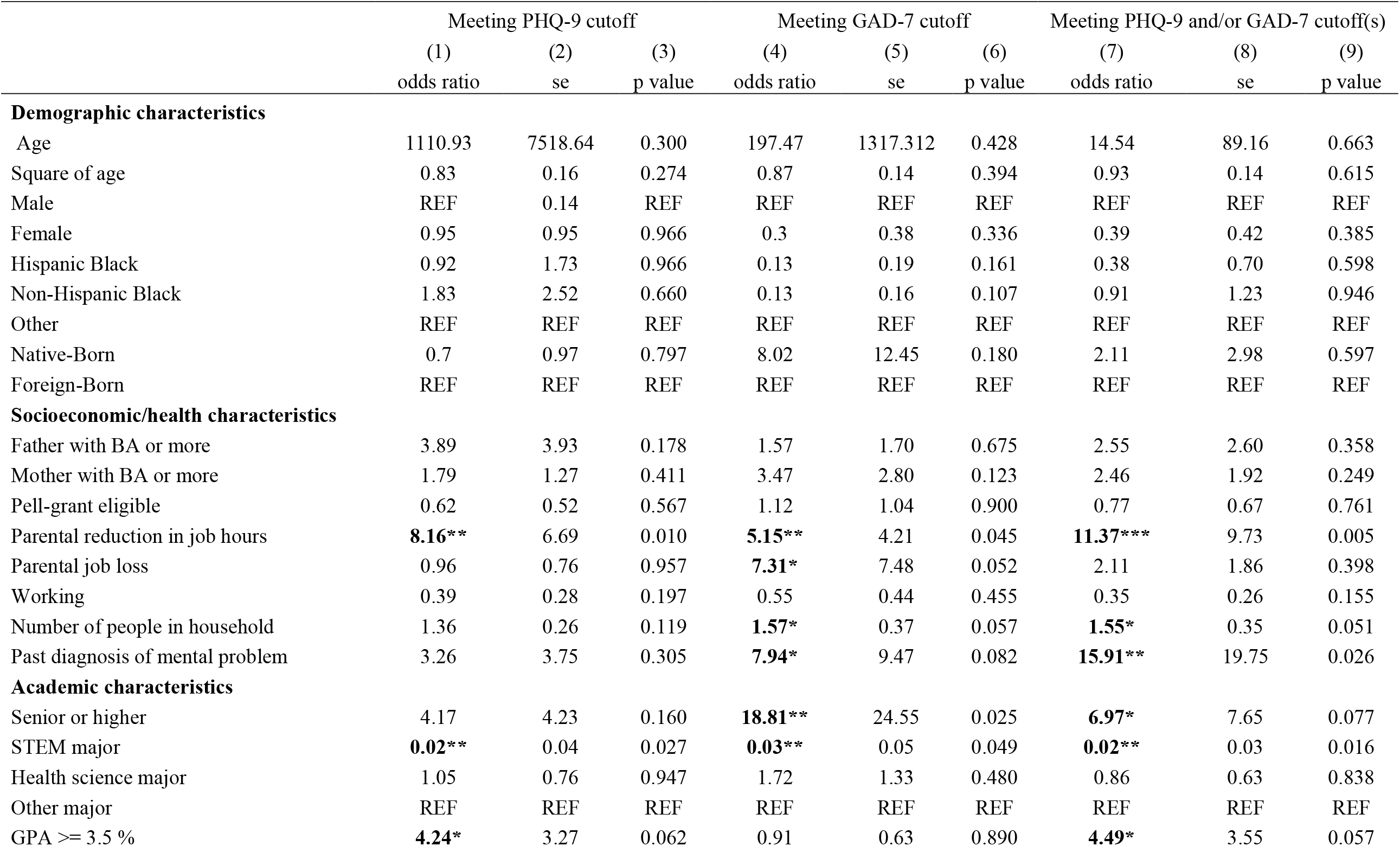
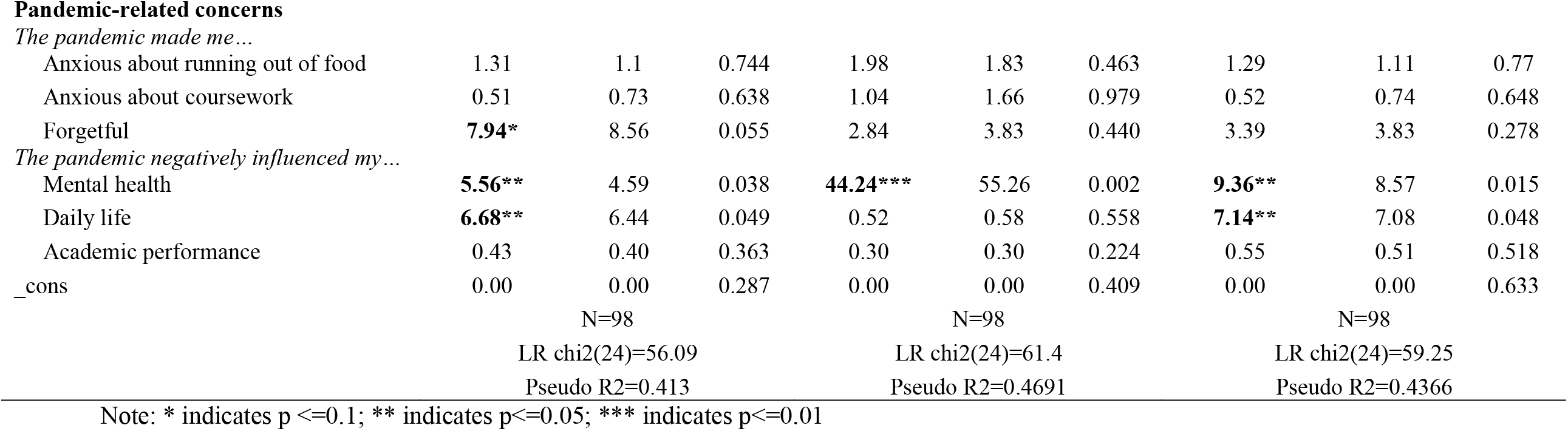
Results from the logistic regression models predicting mental health

Academic characteristics also served as significant predictors of mental health. Being a “senior or higher”, relative to being a non-senior, increased the odds of meeting the GAD-7 cutoff by 18 times and also increased the odds of meeting the GAD-7 cutoff and/or the PHQ-9 GAD-7 cutoff by 7 times, holding all other variables constant. Interestingly, students majoring in STEM were predicted to have lower odds of meeting the cutoffs for PHQ-9 (OR = 0.02, p=0.027), GAD-7 (OR = 0.03, p=0.049), and PHQ-9 and/or GAD-7 (OR=0.02, p=0.016), compared to students majoring in other fields (business, social science, art, or education). Lastly, having a cumulative GPA of 3.5 or higher was predicted to increase the odds of meeting the PHQ-9 cutoff (OR=4.24, p = 0.062) and the odds of meeting the PHQ-9 and/or GAD-7 cutoff(s) (OR=4.49, p =0.057), while holding other variables constant.

In addition, several pandemic-related variables significantly predicted mental health in the present study. Students who reported that the pandemic made them forgetful were 8 times more likely to meet the GAD-7 cutoff compared to those who reported the pandemic did not make them forgetful, holding all other variables constant. Moreover, students who reported the pandemic negatively affected their mental health had much greater odds of meeting the cutoffs for the PHQ-9 (OR=5.56, p=0.038), GAD-7 (OR=44.24, p=0.002), and PHQ-9 and/or GAD-7 (OR=9.36, p=0.015), holding all other variables constant. Finally, students reporting that the pandemic negatively affected their daily life had greater odds of meeting the cutoff for PHQ-9 (OR = 6.68, p =0.049) in addition to PHQ-9 and/or GAD-7 (OR=7.14, p =0.048), holding the covariates constant.

## DISCUSSION AND CONCLUSION

The COVID-19 pandemic has caused a surge in mental health issues around the world, and prior evidence from the United States has suggested disproportionately worse mental health problems among college students as well as racial and ethnic minorities. The present study specifically examined undergraduate students enrolled at an HBCU to determine (1) rates of common mental health issues (i.e., anxiety and depression) and (2) some of the various factors that potentially predict these mental health issues during the pandemic. Our findings indicate notably high rates of anxiety and depression in our sample of HBCU students and that academic characteristics, socioeconomic challenges, and pandemic-related concerns served as significant predictors of these mental health problems.

The present study found that a high rate of HBCU students in our sample met the clinical cutoffs for depression (49%), anxiety (39%), and depression and/or anxiety (52%), using the PHQ-9 and GAD-7 scales to measure depression and anxiety, respectively. Importantly, while these questionnaires do not serve to diagnose psychological disorders, prior research has indicated that those who meet the clinical cutoffs on the PHQ-9 and GAD-7 scales often meet full criteria for depressive or anxiety disorders, respectively, upon further evaluation.

The rates of anxiety and depression observed in our sample were higher than those reported for college students overall during the pandemic. A national survey of college students conducted from mid-March 2020 to May 2020 reported that 41% of college students met the cutoff for depression and 31% met the cutoff for anxiety (Healthy Minds Network & American College Health Association, 2020). In addition, our observed rates of mental health issues were also higher than those observed in a national survey of American adults conducted by the Centers for Disease Control (CDC). For the week of May 14–19, 2020, the CDC reported that 24% of American adults met the cutoff for depression, 28% met the cutoff for anxiety, and 34% met the cutoff for depression and/or anxiety. Also, in the CDC survey, non-Hispanic Black Americans had higher rates of mental health issues relative to Americans overall, but the numbers were still lower than those observed in our sample of HBCU students. For the week of May 14–19, 2020, the CDC reported that 29% of non-Hispanic Black Americans met the cutoff for depression, 33% met the cutoff for anxiety, and 39% met the cutoff for depression and/or anxiety.

One possible interpretation of these finding is that Black/African American college students may have experienced greater mental health distress as a result of the pandemic compared to college students overall as well as Black/African Americans overall. This may be attributed, in part, to the combination of concerns faced by college students and racial and ethnic minorities during the pandemic. Importantly, prior research has indicated that attending an HBCU has a positive effect on mental health of Black students, partly because the HBCU environment instills a greater sense of belonging and less marginalization and discrimination (Watkins et al., 2007; Mushonga & Henneberger,, 2020). Therefore, it is possible that Black students attending a predominantly white institution (PWI) might demonstrate worse mental health measures during the pandemic compared to our sample of HBCU students.

Regarding our analysis of the various factors that potentially influenced mental health in our sample, it was surprising that demographic characteristics did not play a significant role. There was no effect of age, sex, or place of birth on the odds of meeting the clinical cutoff for depression or anxiety. In addition, race/ethnicity did not significantly influence mental health measures in our sample. Specifically, Hispanic and non-Hispanic Black students did not show differences in the likelihood of meeting the clinical cutoffs compared to non-Black students. One prominent reason for this observation may be the fact that the small group of non-Black students in our sample was mostly comprised of other racial and ethnic minorities who may experience similar mental health challenges as a result of the pandemic.

Among the factors shown to impact mental health in our study were socioeconomic challenges. In particular, students whose parents experienced a cut in hours during the pandemic were more likely to meet the clinical cutoffs for depression, anxiety, and depression and/or anxiety. Similarly, students whose parents lost their job during the pandemic were more likely to meet the cutoff for anxiety. Prior research has shown that stress resulting from parental job loss has the potential to trickle down and negatively impact their children’s mental health (Schaller & Zerpa, 2019). Specifically, it is possible that, for the students in our sample, parental job loss and cuts in hours may have negatively impacted family finances and made it more difficult for the student to focus on their studies and to pay for their college tuition, which has the potential to harm mental health.

Importantly, the finding that job loss increased mental health issues in our sample is noteworthy considering that Black/African Americans have faced greater unemployment and slower job recovery during the pandemic, compared to non-Hispanic White Americans (Sáenz & Sparks, 2020). The RAND organization reported that in April 2020 unemployment rate was above 15% for Black workers and below 15% for White workers (Williams, 2020). By August 2020, the unemployment rate decreased considerably to 7% for White workers, whereas it remained high among Black workers (about 13%; Williams, 2020; <u>link</u>). Our present findings suggest that greater unemployment among Black Americans may lead to a higher proportion of mental health problems for this demographic group during the pandemic. However, further research will be required to determine whether the influence of job loss and reduction in work hours on mental health of Black Americans persists on a national level.

Interestingly, the present study also found that the number of people living in the household had a negative effect on the students’ mental health, increasing the likelihood of anxiety, as well as depression and/or anxiety. Importantly, when the survey was conducted in May 2020, students were not permitted on campus due to COVID restrictions and had to complete their courses via remote/virtual learning. Staying at home may have increased mental health issues in part due to limited space and resources, and previous research has shown that living in a household with many occupants has a negative effect on mental health and general well-being (Clapham, Foye, and Christian, 2018). Moreover, a large household may increase the likelihood of virus transmission, especially if the household includes family members working high-risk essential jobs (Martin et al., 2020). Prior research has indicated that Black/African Americans are more likely to contract the COVID-19 virus and face worse health outcomes after infection, compared to non-Hispanic Whites (Oppel Jr, Gebeloff, Lai, Wright, & Smith, 2020).

Student’s academic characteristics also played a significant role in whether they met the criteria for mental health issues. Being a senior or having a good GPA (3.5 or higher) were each associated with greater likelihood of anxiety and depression. The COVID-19 pandemic has crippled the job market, potentially making it more difficult for graduating seniors to find available jobs (Aucejo, French, Araya, & Zafar, 2020), whereas other students (freshmen, sophomores, and juniors) may not be as deeply affected by the difficult job market. The observation that students with high GPAs face greater mental health distress may be due to the quick, nonvoluntary transition to online learning during the pandemic, which has proven stressful for college students (Liu, Pinder-Amaker, Hahm, & Chen, 2020; Wang et al., 2020). The present findings also indicated that majoring in a STEM field was associated with lower likelihood of depression, anxiety, and depression and/or anxiety. This finding is largely consistent with prior research showing that students majoring in non-STEM fields, such as humanities or the arts, demonstrate worse mental health outcomes than their peers (Lipson, Zhou, Wagner, Beck, & Eisenberg, 2016).

In sum, the present study demonstrated high rates of mental health issues among HBCU students during the COVID-19 pandemic, and that mental health problems may be predicted based on the student’s academic characteristics, socioeconomic challenges, and pandemic-related concerns. The study also raised new questions that may be explored in future research. For instance, it should be noted that our study only examined mental health of HBCU students from a single university. Further studies will be required to confirm that the high rates of mental health issues observed in our sample would be reflected in a national survey of Black/African American college students. In addition, the present survey only included mental health measures for depression and anxiety. Future research would benefit from examining additional mental health issues common among college students and Black/African Americans, such as eating disorders and post-traumatic stress (Abrams, Allen, & Gray, 1993; Fitzpatrick & Boldizar, 1993; Eisenberg, Nicklett, Roeder, & Kirz, 2011).

## Data Availability

Data available upon request.

